# A Fully Automatic Deep Learning System for COVID-19 Diagnostic and Prognostic Analysis

**DOI:** 10.1101/2020.03.24.20042317

**Authors:** Shuo Wang, Yunfei Zha, Weimin Li, Qingxia Wu, Xiaohu Li, Meng Niu, Meiyun Wang, Xiaoming Qiu, Hongjun Li, He Yu, Wei Gong, Yan Bai, Li Li, Yongbei Zhu, Liusu Wang, Jie Tian

## Abstract

Coronavirus disease 2019 (COVID-19) has spread globally, and medical resources become insufficient in many regions. Fast diagnosis of COVID-19, and finding high-risk patients with worse prognosis for early prevention and medical resources optimization is important. Here, we proposed a fully automatic deep learning system for COVID-19 diagnostic and prognostic analysis by routinely used computed tomography.

We retrospectively collected 5372 patients with computed tomography images from 7 cities or provinces. Firstly, 4106 patients with computed tomography images and gene information were used to pre-train the DL system, making it learn lung features. Afterwards, 1266 patients (924 with COVID-19, and 471 had follow-up for 5+ days; 342 with other pneumonia) from 6 cities or provinces were enrolled to train and externally validate the performance of the deep learning system.

In the 4 external validation sets, the deep learning system achieved good performance in identifying COVID-19 from other pneumonia (AUC=0.87 and 0.88) and viral pneumonia (AUC=0.86). Moreover, the deep learning system succeeded to stratify patients into high-risk and low-risk groups whose hospital-stay time have significant difference (p=0.013 and 0.014). Without human-assistance, the deep learning system automatically focused on abnormal areas that showed consistent characteristics with reported radiological findings.

Deep learning provides a convenient tool for fast screening COVID-19 and finding potential high-risk patients, which may be helpful for medical resource optimization and early prevention before patients show severe symptoms.

**Take-home message:** Fully automatic deep learning system provides a convenient method for COVID-19 diagnostic and prognostic analysis, which can help COVID-19 screening and finding potential high-risk patients with worse prognosis.

## Introduction

In Dec. 2019, the novel coronavirus disease 2019 (COVID-19) occurred in Wuhan, China and became a global health emergency very fast with more than 170,000 people infected [1-3]. Due to its high infection rate, fast diagnosis and optimized medical resource assignment in epidemic areas are urgent. Accurate and fast diagnosis of COVID-19 can help isolating infected patients to slow the spread of this disease. On the other hand, in epidemic area, insufficient medical resources have become a big challenge [4]. Therefore, finding high-risk patients with worse prognosis for prior medical resources and special care is crucial in the treatment of COVID-19.

Currently, reverse transcription polymerase chain reaction (RT-PCR) is used as the gold truth for diagnosing COVID-19. However, the limited sensitivity of RT-PCR and the shortage of testing kits in epidemic areas increase the screening burden, and many infected people are thereby not isolated immediately [5, 6]. This accelerates the spread of COVID-19. On the other hand, due to the lack of medical resources, many infected patients cannot receive immediate treatment. In this situation, finding high-risk patients with worse prognosis for prior treatment and early prevention is important. Consequently, fast diagnosis, finding high-risk patients with worse prognosis are very helpful for the control and management of COVID-19.

In recent studies, radiological findings demonstrated that computed tomography (CT) has great diagnostic and prognostic value for COVID-19. For example, CT showed much higher sensitivity than RT-PCR in diagnosing COVID-19 [5, 6]. For patients with COVID-19, bilateral lung lesions consisting of ground-glass opacities (GGO) were frequently observed in CT images [6-8]. Even in asymptomatic patients, abnormalities and changes were observed in serial CT [9, 10]. As a common diagnostic tool, CT is easy and fast to acquire without adding much cost. Building a sensitive diagnostic tool using CT image can accelerate the diagnostic process and is complementary to RT-PCR. On the other hand, predicting personalized prognosis using CT image can identify the potential high-risk patients who are more likely to become severe and need urgent medical resources.

Deep learning (DL) as an artificial intelligence method, has shown promising results in assisting lung disease analysis using CT images [11-14]. Benefiting from the strong feature learning ability, DL can mine features that are related to clinical outcomes from CT images automatically. Features learned by DL models can reflect high-dimensional abstract mappings which are difficult for human to sense but are strongly associated with clinical outcomes. Different from the published DL models [15, 16], we aim to provide a fully automatic DL system for COVID-19 diagnostic and prognostic analysis. Without requiring any human-assisted annotation, this novel DL system is fast and robust in clinical use. Moreover, we collected a large multi-regional dataset for training and validating the proposed DL system, including 1266 patients (471 had follow-up) from six cities or provinces. Notably, different with many studies using transfer learning from natural images. We collected a large auxiliary dataset including 4106 patients with chest CT images and gene information to pre-train the DL system, aiming at making the DL system learn lung features that can reflect the association between micro-level lung functional abnormalities and chest CT images.

## Methods

### Study design and participants

The institutional review board of the seven hospitals (supplementary methods 1) approved this multi-regional retrospective study and waived the need to obtain informed consent from the patients. In this study, we collected two datasets: COVID-19 dataset (n=1266) and CT-EGFR dataset (n=4106). In the COVID-19 dataset, 1266 patients were finally included who met the following inclusion criteria: (i) RT-PCR confirmed COVID-19; (ii) lab-confirmed other types of pneumonia before Dec. 2019; (iii) have non-contrast enhanced chest CT at diagnosis time. Since RT-PCR has a relatively high false-negative rate, we collected other types of pneumonia before Dec. 2019 when the COVID-19 did not show up to guarantee the diagnosis of typical pneumonia are correct. In the COVID-19 dataset, patients from Wuhan city and Henan province formed the training set; patients from Anhui province formed the external validation set 1; patients from Heilongjiang province formed the validation set 2; patients from Beijing formed the validation set 3; patients from Huangshi city formed the validation set 4 (figure 1).

**Figure 1.**
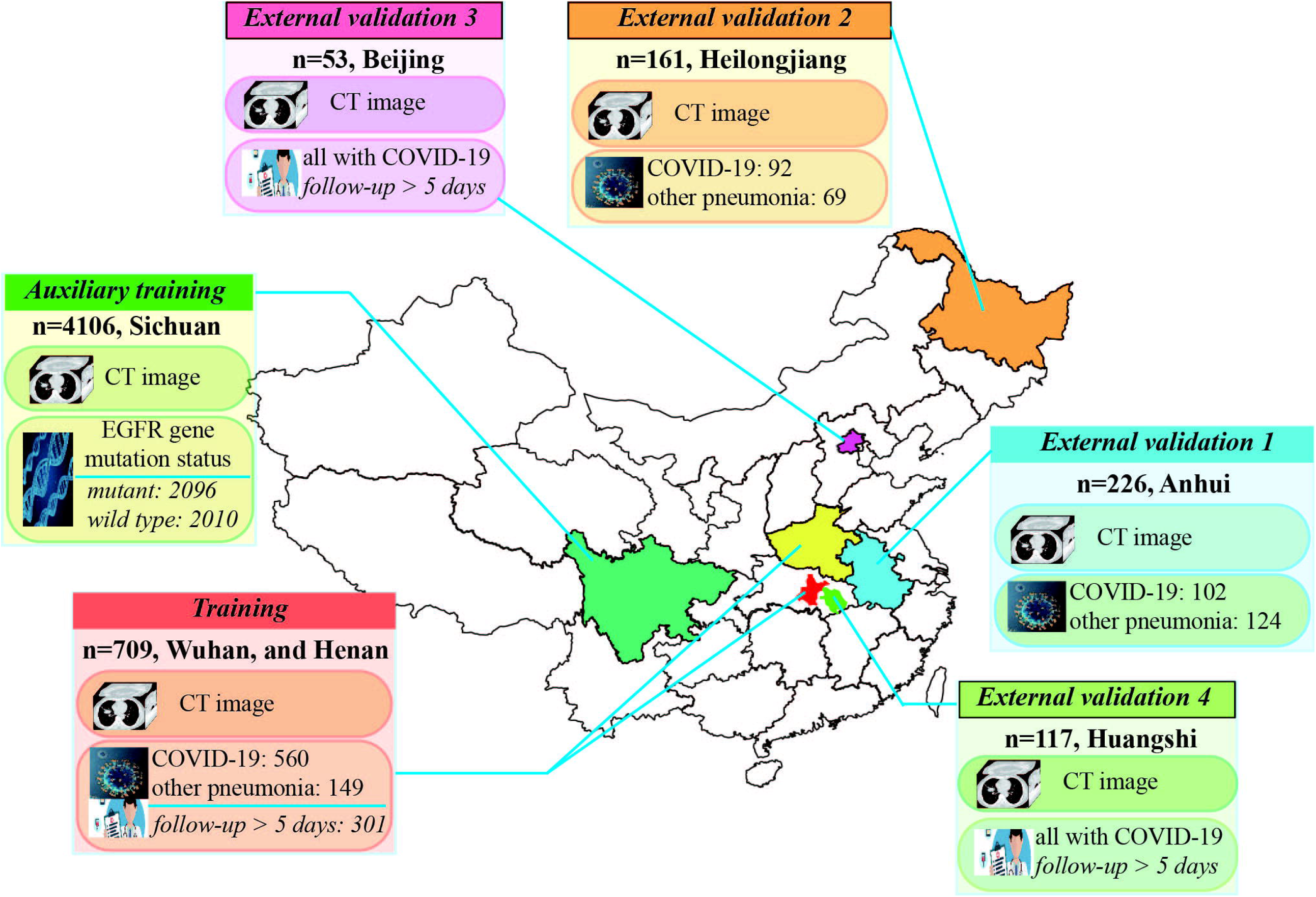
Datasets used in this study. A total of 5372 patients with CT images from 7 cities or provinces were enrolled in this study. The auxiliary training set includes 4106 patients with lung cancer and EGFR gene mutation status information, and is used to pre-train the COVID-19Net to learn lung features from CT images. The training set includes 709 patients from Wuhan city and Henan province. The external validation set 1 (226 patients) from Anhui province, and the external validation set 2 (161 patients) from Heilongjiang province are used to test the diagnostic performance of the DL system. The external validation set 3 (53 patients with COVID-19) from Beijing, and the external validation set 4 (117 patients with COVID-19) from Huangshi city are used to evaluate the prognostic performance of the DL system.

In the CT-EGFR dataset, 4106 patients with lung cancer were finally included who met the following criteria: (i) epidermal growth factor receptor (EGFR) gene sequencing was obtained; (ii) non-contrast enhanced chest CT data obtained within 4 weeks before EGFR gene sequencing. The CT-EGFR dataset was used for auxiliary training of the DL system, making the DL system learn lung features automatically. CT scanning parameters about the COVID-19 and CT-EGFR datasets were available in supplementary methods S1.

For prognostic analysis, 471 patients with COVID-19 and regular follow-up for at least 5 days were used. We defined the prognostic end event as the hospital-stay time which is counted from the diagnosis of COVID-19 to the time when the patient is allowed to discharge hospital (supplementary methods S2). A short hospital-stay time corresponds to good prognosis, and a long hospital-stay time means worse prognosis. Patients with long hospital-stay time take longer time to recover, and are defined as high-risk patients in this study. These patients need prior medical resources and special care since they are more likely to become severe.

The training set was used to train the proposed DL system; the validation set 1 and 2 were used to evaluate the diagnostic performance of the DL system; and the validation set 3 and 4 were used for evaluating the prognostic performance of the DL system.

### The fully automatic deep learning system for COVID-19 diagnostic and prognostic analysis

The proposed DL system includes three parts: automatic lung segmentation, non-lung area suppression, and COVID-19 diagnostic and prognostic analysis. In this DL system, two DL networks were involved: DenseNet121-FPN for lung segmentation in chest CT image, and the proposed novel COVID-19Net for COVID-19 diagnostic and prognostic analysis. DL is a family of hierarchical neural networks that aim at learning the abstract mapping between raw data to the desired clinical outcome. The computational units in DL model are defined as layers and they are integrated to simulate the inference process of human brain. The main computational formulas are convolution, pooling, activation and batch normalization as defined in the supplementary methods S3.

### Automatic lung segmentation

Routinely used chest CT image includes some non-lung areas (muscle, heart, et al.) and blank space outside body. To focus on analyzing lung area, we used a fully automatic DL model (DenseNet121-FPN) [17, 18] to segment lung areas in chest CT image. This model is pre-trained using 1.4 million natural images, and fine-tuned on VESSEL12 dataset [19] (supplementary methods S4).

Through this automatic lung segmentation procedure, we acquired the lung mask in CT image. However, some inflammatory tissues attaching to lung wall may be excluded falsely by the DenseNet121-FPN model. To increase the robustness of the DL system, we used the cubic bounding box of the segmented lung mask to crop lung areas in CT image, and defined this cubic lung area as lung-ROI (figure 2). In this lung-ROI, all inflammatory tissues and the whole lung were correctly reserved, and most areas outside of lung were eliminated.

**Figure 2.**
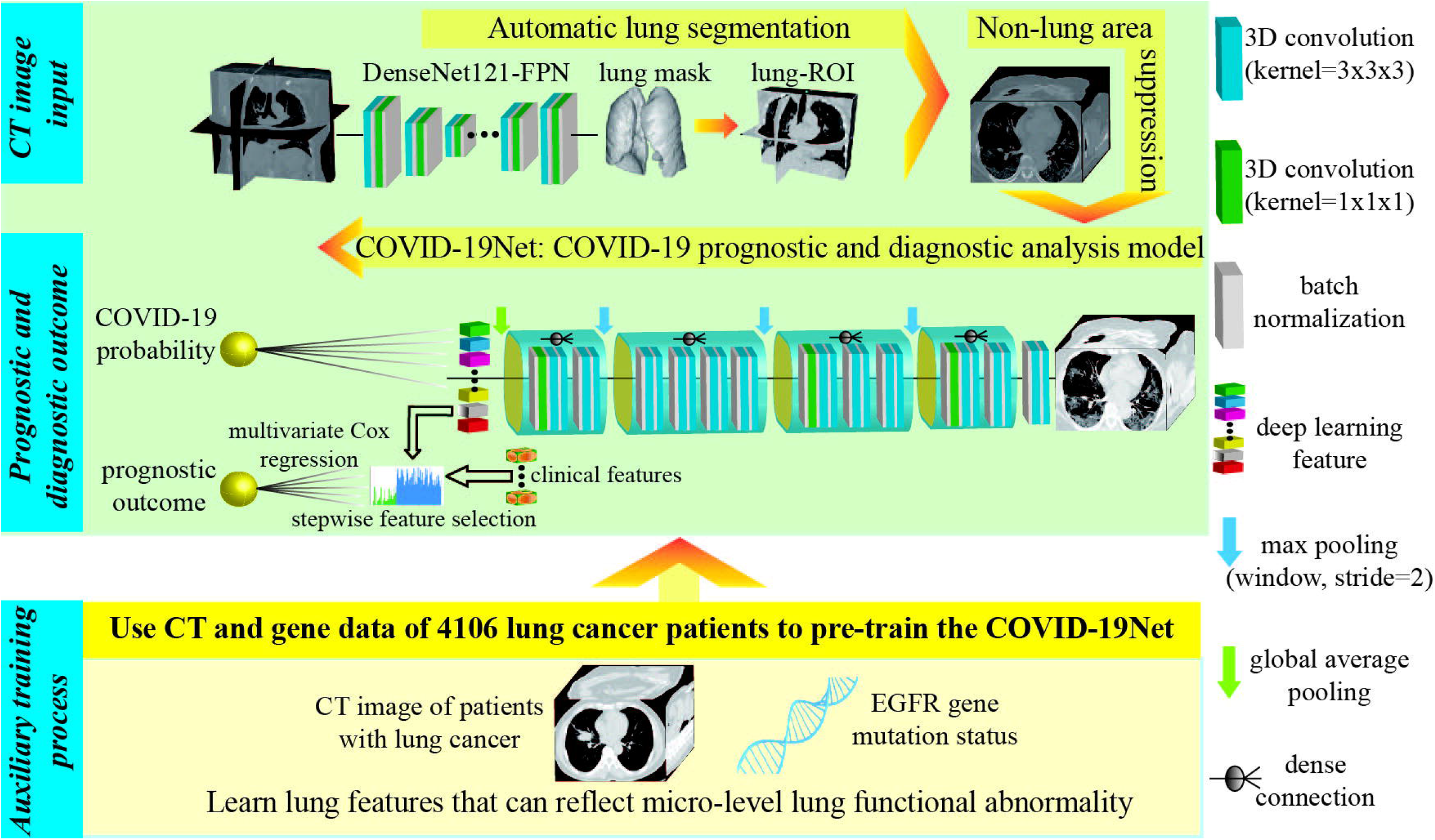
Illustration of the proposed DL system. Given the chest CT scanning of a patient, the DL system predicts the probability of the patient has COVID-19 and the prognosis of this patient directly without any human-annotation. The DL system includes three parts: automatic lung segmentation (DenseNet121-FPN), non-lung area suppression, and COVID-19 diagnostic and prognostic analysis (COVID-19Net). To let the COVID-19Net learn lung features from large dataset, we used the auxiliary training process for pre-training, which trained the DL network to predict EGFR gene mutation status using CT images of 4106 patients. The dense connection in this figure means each convolutional layer is connected to all of its previous convolutional layers inside the same dense block.

### Non-long area suppression

After the above processing, some non-lung tissues or organs (e.g., spine, heart) inside the lung-ROI may also exists. Consequently, we proposed a non-lung area suppression operation to suppress the intensities of non-lung areas inside the lung-ROI (supplementary methods S4). Finally, the lung-ROI is standardized by z-score normalization, and resized to the size of 48×240×360 for further process.

### Deep learning model for COVID-19 diagnosis and prognosis

After non-lung area suppression operation, the standardized lung-ROI is sent into the COVID-19Net for diagnostic and prognostic analysis. In figure 2, we illustrated the topological structure of the proposed novel COVID-19Net (supplementary table S1). This DL model used DenseNet-like structure [17], consisting of four dense blocks, where each dense block is multiple stacks of convolution, batch normalization, and ReLU activation layers. Inside each dense block, we used dense connection to consider multi-level image information. At the end of the last convolutional layer, we used global average pooling to generate the 64-dimensional DL features. Finally, the output neuron is fully connected to the DL features to predict the probability of the input patient has COVID-19.

To enable the COVID-19Net learn discriminative features associated with COVID-19, a large training set is needed. Consequently, we proposed a two-step transfer learning process. Firstly, we proposed an auxiliary training process use our collected large CT-EGFR dataset (4106 patients) as illustrated in figure 2. In this auxiliary training process, we trained the COVID-19Net to predict EGFR mutation status (EGFR-mutant or EGFR wild type) use the lung-ROI [11]. Benefitting from the large CT-EGFR dataset, the COVID-19Net learned CT features that can reflect the associations between micro-level lung functional abnormality and macro-level CT images.

In the second training process, we transferred the pre-trained COVID-19Net to the COVID-19 dataset to specifically mine lung characteristics associated with COVID-19. After iterative training process in the COVID-19 dataset (supplementary methods S5), the COVID-19Net can predict a probability of the input patient being infected with COVID-19; this probability was defined as DL score in this study.

To predict personalized prognosis for patient with COVID-19, we extracted the 64-dimensional DL feature from the COVID-19Net to build a prognostic prediction model. Firstly, we combined the 64-dimensional DL feature and clinical features (age, sex, and comorbidity) to construct a combined feature vector. Afterwards, we used stepwise method to select prognostic features. These selected features were then used to build a multivariate Cox proportional hazard (CPH) model to predict the hazard of the patient needing a long hospital-stay time to recover.

### Visualization of lung features learned by the DL system

Through the two-step transfer learning technique, the DL system learned lung features from CT images of 4815 patients. To further understand the inference process of the DL system, we used DL visualization algorithm to analyze features learned by the COVID-19Net from two perspectives: 1) visualizing DL-discovered suspicious lung area that contribute most for identifying COVID-19 for the DL system; 2) visualizing the feature patterns extracted by hierarchical convolutional layers in the COVID-19Net (supplementary methods S6, S7).

### Statistical analysis

Area under the receiver operating characteristic (ROC) curve (AUC), accuracy, sensitivity, and specificity were used to assess the performance of the DL system in diagnosing COVID-19. Kaplan-Meier analysis and log-rank test were used to evaluate the performance of the DL system for prognostic analysis. The implementation of the DL system used the Keras 2.0.0 toolkit and Python 2.7.

## Results

Clinical characteristics of patients in the COVID-19 dataset were presented in table 1. This dataset was collected from six cities or provinces including Wuhan in China.

**Table 1.**
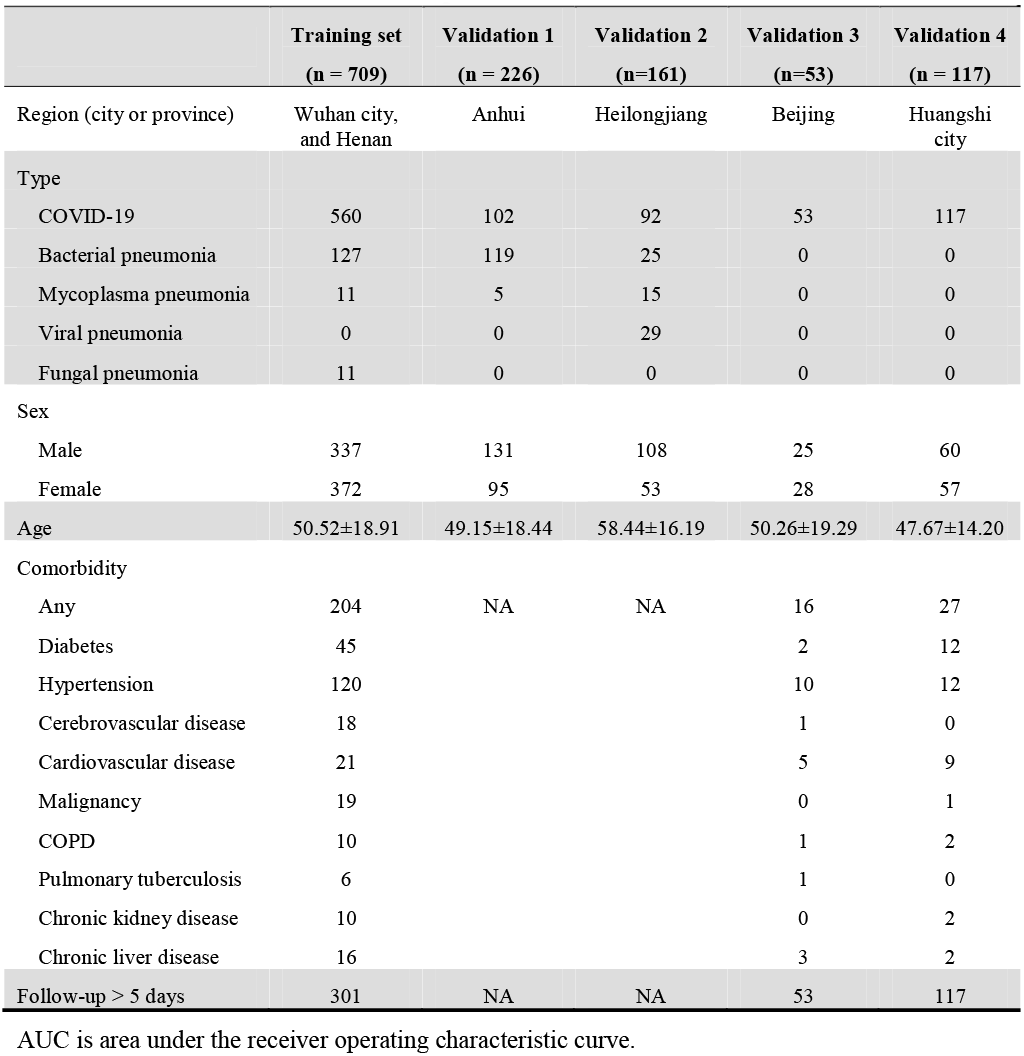
Clinical characteristics of patients.

|  | Training set<br>(n = 709) | Validation 1<br>(n = 226) | Validation 2<br>(n=161) | Validation 3<br>(n=53) | Validation 4<br>(n = 117) |
| --- | --- | --- | --- | --- | --- |
| Region (city or province) | Wuhan city,<br>and Henan | Anhui | Heilongjiang | Beijing | Huangshi<br>city |
| Type |  |  |  |  |  |
| COVID-19 | 560 | 102 | 92 | 53 | 117 |
| Bacterial pneumonia | 127 | 119 | 25 | 0 | 0 |
| Mycoplasma pneumonia | 11 | 5 | 15 | 0 | 0 |
| Viral pneumonia | 0 | 0 | 29 | 0 | 0 |
| Fungal pneumonia | 11 | 0 | 0 | 0 | 0 |
| Sex |  |  |  |  |  |
| Male | 337 | 131 | 108 | 25 | 60 |
| Female | 372 | 95 | 53 | 28 | 57 |
| Age | 50.52±18.91 | 49.15±18.44 | 58.44±16.19 | 50.26±19.29 | 47.67±14.20 |
| Comorbidity |  |  |  |  |  |
| Any | 204 | NA | NA | 16 | 27 |
| Diabetes | 45 |  |  | 2 | 12 |
| Hypertension | 120 |  |  | 10 | 12 |
| Cerebrovascular disease | 18 |  |  | 1 | 0 |
| Cardiovascular disease | 21 |  |  | 5 | 9 |
| Malignancy | 19 |  |  | 0 | 1 |
| COPD | 10 |  |  | 1 | 2 |
| Pulmonary tuberculosis | 6 |  |  | 1 | 0 |
| Chronic kidney disease | 10 |  |  | 0 | 2 |
| Chronic liver disease | 16 |  |  | 3 | 2 |
| Follow-up > 5 days | 301 | NA | NA | 53 | 117 |

3 AUC is area under the receiver operating characteristic curve.

### Diagnostic performance of the DL system

Table 2 and figure 3 illustrated the diagnostic performance of the DL system. In the training set, the DL system showed good diagnostic performance (AUC=0.90). This performance was further confirmed in the two external validation sets (AUC=0.87 and 0.88). The good performance in the validation cohorts indicated that the DL system generalized well on diagnosing COVID-19 of unseen new patients. Meanwhile, we illustrated the ROC curves of the DL system in the three sets in figure 3a. The DL score revealed a significant difference between COVID-19 and other pneumonia groups in the three sets (p<0.0001).

**Table 2.** Diagnostic performance of the DL system.

|  | **Training**<br>**(n=709)** | **Validation 1**<br>**(n=226)** | **Validation 2**<br>**(n=161)** | **Validation 2-viral**<br>**(n=120)** |
| --- | --- | --- | --- | --- |
| **AUC (95%CI)** | 0.90 (0.89-0.91) | 0.87 (0.86-0.89) | 0.88 (0.86-0.90) | 0.86 (0.83, 0.89) |
| **Accuracy (%)** | 81.24 | 78.32 | 80.12 | 85.00 |
| **Sensitivity (%)** | 78.93 | 80.39 | 79.35 | 79.35 |
| **Specificity (%)** | 89.93 | 76.61 | 81.16 | 71.43 |

2 AUC is area under the receiver operating characteristic curve.

**Figure 3.**
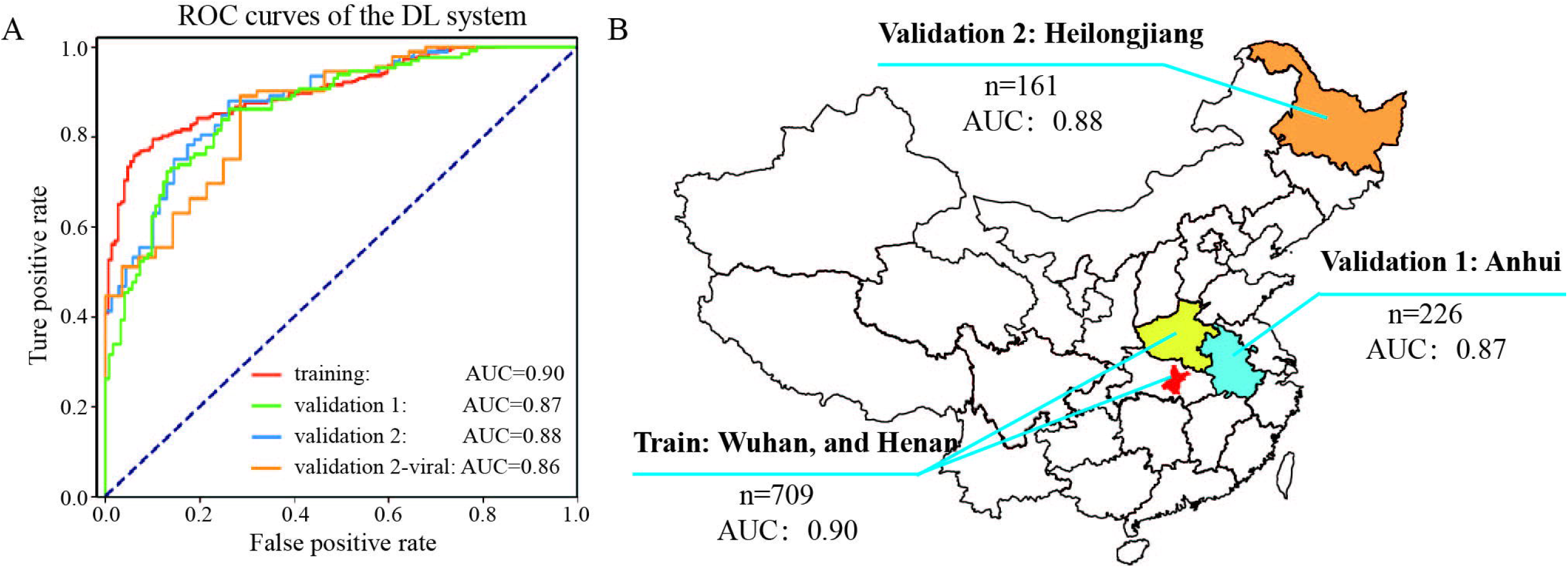
Diagnostic performance of the DL system. **a)**. ROC curves of the DL system in the training set and the two independent external validation sets. Validation 2-viral is a stratified analysis using the patients with COVID-19 and viral pneumonia in the validation set 2. **b)**. AUC and distribution of the training set and the two external validation datasets.

In other types of pneumonia, viral pneumonia has similar radiological characteristics to COVID-19, and therefore is more difficult to identify. Consequently, we performed a stratified analysis in the validation set 2. Table 1 indicated that the DL system also achieved good results in distinguish COVID-19 to other viral pneumonia (AUC=0.86).

### Prognostic performance of the DL system

In the COVID-19 dataset, 471 patients had follow-up for 5+ days. Through the stepwise prognostic feature selection, 3 features were selected (supplementary table S2). These selected prognostic features were fed into the multivariate CPH model to predict a hazard value for each patient. We used median value of the hazards in the training set as cut-off value to stratify patients into high-risk and low-risk groups. This cut-off value was also applied to the validation set 3 and 4. Kaplan-Meier analysis in figure 4 demonstrated that patients in high-risk and low-risk groups had significant difference in hospital-stay time in the three datasets (p<0.0001, p=0.013, and p=0.014, log-rank test).

**Figure 4.**
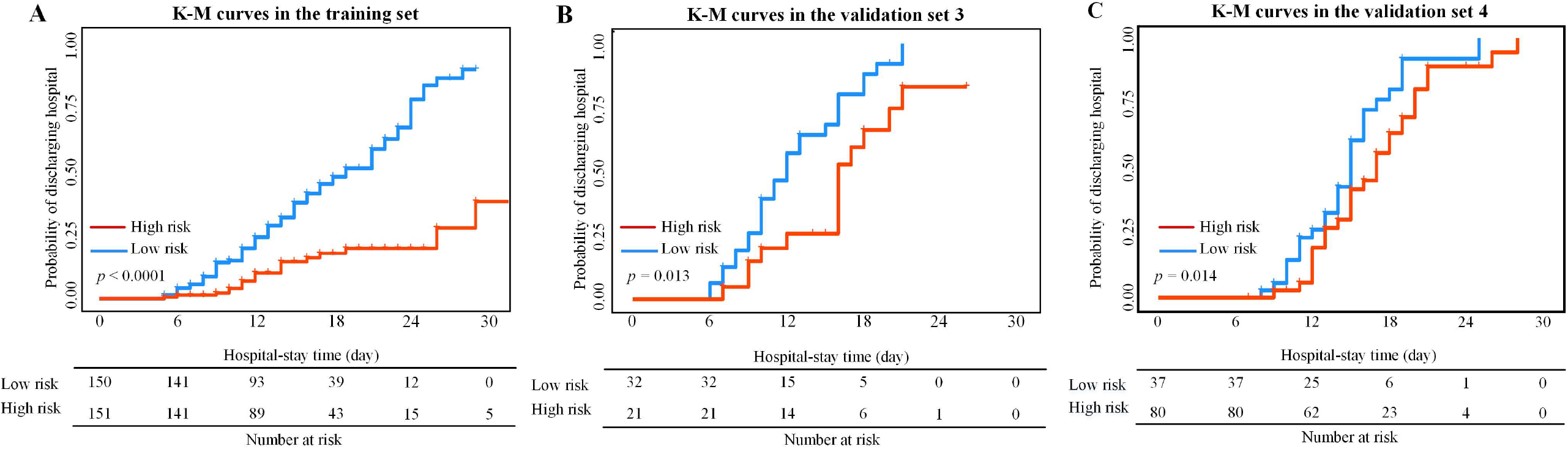
Kaplan-Meier analysis of the prognostic performance of the DL system. Vertical lines in this figure represents censored data.

For a given patient, if the DL system predicts him/her as COVID-19, the DL system predicts a prognostic hazard simultaneously. For the patients who are predicted as high-risk by the DL system, prior medical resources and special care are suggested.

### Suspicious lung area discovered by the DL system

Through DL visualization algorithm [20, 21], we are able to visualize the lung area that draws most attention to the DL system. These DL-discovered suspicious lung areas usually demonstrated abnormal characteristics consistent with radiologists’ findings. Figure 5 illustrated DL-discovered suspicious lung areas of eight patients with COVID-19. From this figure, we can see that although the input lung-ROI to the DL system includes some non-lung tissues such as muscle and bones, the DL system can always focus on areas inside lung for prediction instead of being disturbed by other tissues.

**Figure 5.**
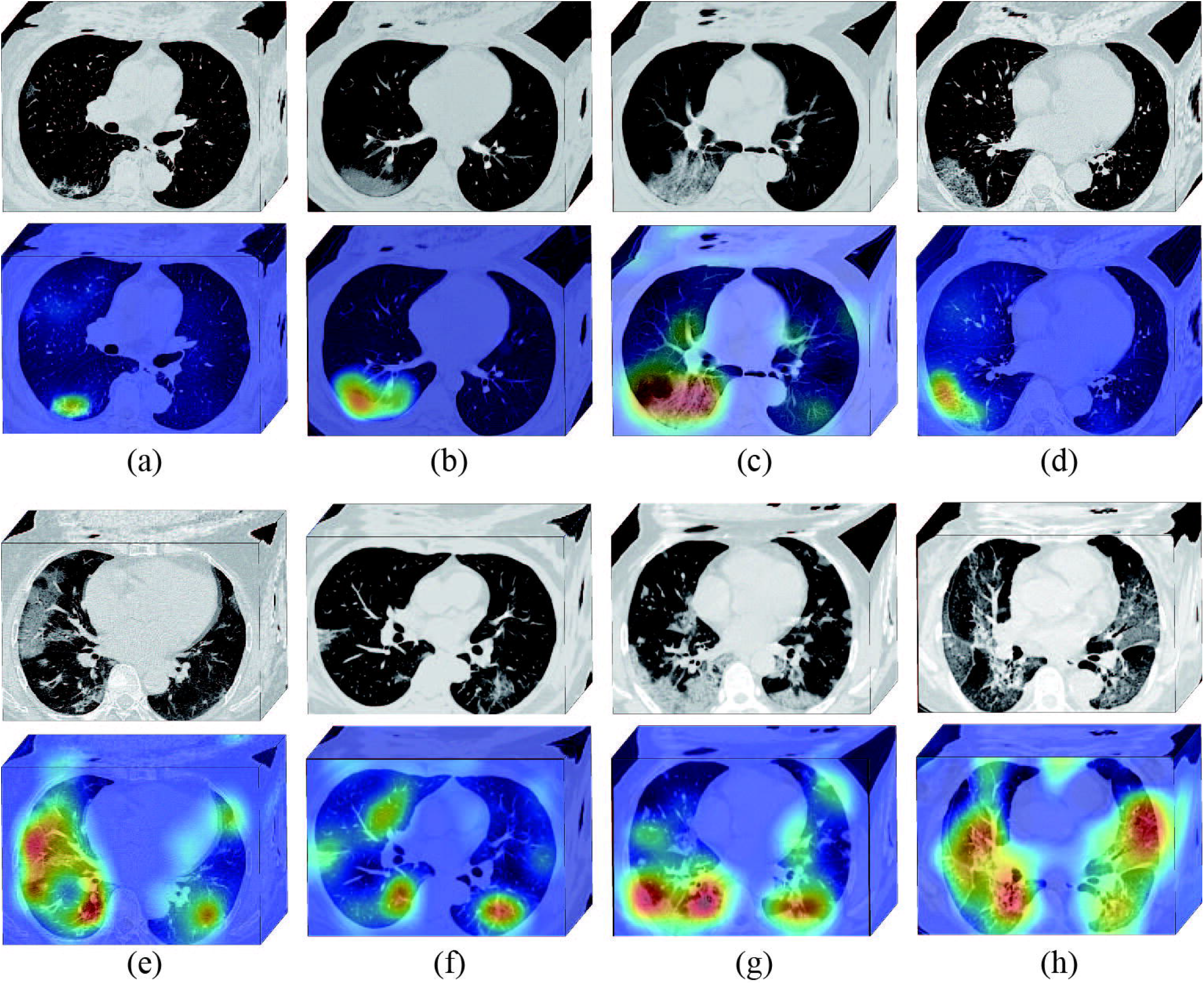
DL-discovered suspicious lung area. **(a)-(h)** are CT images of eight patients with COVID-19. The first and the third rows are CT images of the patients (these CT images are processed by the DL system). The second and the fourth rows are heat maps of the DL-discovered suspicious lung area. In the heat map, areas with bright red color are more important than dark blue areas.

Moreover, the DL-discovered suspicious lung areas showed high overlap with the actual inflammatory areas. In figure 5 a-d, we can see that, although we did not involve any human-annotation in the DL system, the DL system focused on the GGO area automatically for inference. This is consistent with radiologists’ experiences that many COVID-19 illustrated GGO features [6, 9]. In figure 5 e-h, the DL-discovered suspicious lung areas distributed on bilateral lung, and mainly focused on lesions with consolidation, GGO, diffuse or mixture patterns. When comparing these DL-discovered suspicious lung areas with actual abnormal lung areas, we found a high overlap and consistent.

Although we did not use human annotation (e.g., human annotated ROI) to tell the DL system where to watch, the DL system is capable of discovering the abnormal and important lung areas automatically. This phenomenon could come from the advantage of using the large CT-EGFR dataset and the large COVID-19 dataset for training.

### DL feature visualization

Since DL is an end-to-end prediction model that learns abstract mappings between lung CT image and COVID-19 directly, it is helpful to explain the inference process of the DL system. The most important component of DL model is convolutional filter. Therefore, we visualized the 3-dimensional feature patterns extracted by hierarchical convolutional layers in figure 6. The shallow convolutional layer learned low-level simple features such as spindle edges (figure 6a) and wave-like edges (figure 6b). A deeper convolutional layer learned more complex and detailed features (figure 6c). When going deeper, the feature pattern became more abstract and lack visual characteristics (figure 6d) for our eyes. However, these high-level feature patterns are more related to COVID-19 information.

**Figure 6.**
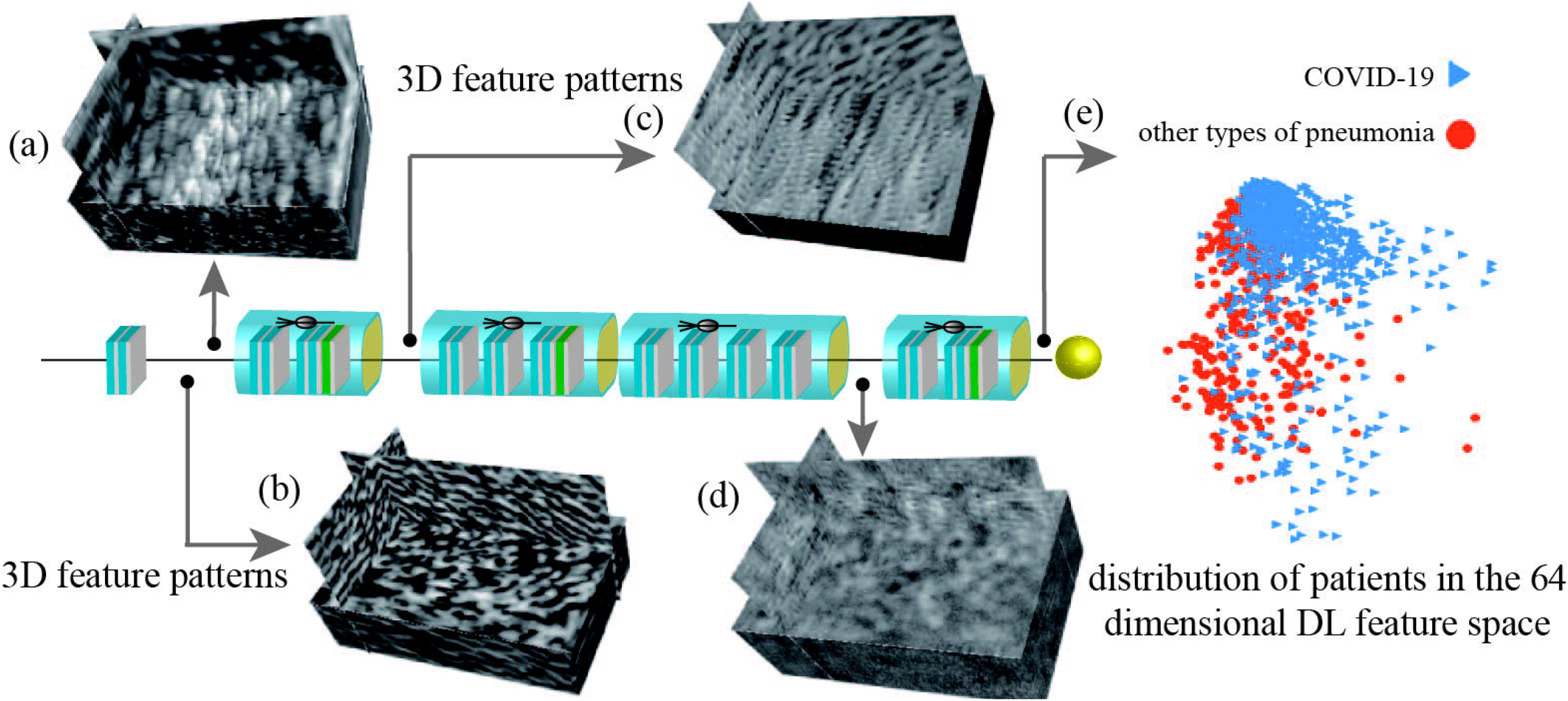
DL feature visualization. **(a)-(d)** are four 3-dimensional (3D) convolutional filters from different convolutional layers. **(e)** is the distribution of patients in the 64-dimensional DL feature space. For display convenience, the 64-dimensional DL feature space is reduced to 2-dimensional by principle component analysis algorithm.

At the end of the DL model, the outputs of convolutional filters were compressed into a 64-dimensional vector, which was defined as DL feature. In figure 6e, we reduced the 64-dimensional DL feature into two-dimensional space to see the DL feature distribution in the two classes (COVID-19 vs. other types of pneumonia). This figure demonstrated that the two classes distributed separately in the DL feature space, which means the DL features are discriminative to identify COVID-19 from other types of pneumonia.

## Discussion

In this study, we proposed a novel fully automatic DL system using raw chest CT image to help COVID-19 diagnostic and prognostic analysis. To let the DL system mine lung features automatically without involving any time-consuming human annotation, we used a two-step transfer learning strategy. Firstly, we collected 4106 lung cancer patients with both CT image and EGFR gene sequencing. Through training in this large CT-EGFR dataset, the DL system learned hierarchical lung features that can reflect the associations between chest CT image and micro-level lung functional abnormality. Afterwards, we collected a large multi-regional COVID-19 dataset (n=1266) from 6 cities or provinces to train and validate the diagnostic and prognostic performance of the DL system.

The good diagnostic and prognostic performance of the DL system illustrates that DL could be helpful in the epidemic control of COVID-19 without adding much cost. Given a suspected patient, CT scanning can be acquired within minutes. Afterwards, this DL system can be applied to predict the probability of the patient has COVID-19. If the patient is diagnosed as COVID-19, the DL system also predicts his/her prognostic situation simultaneously, which can be used to find potential high-risk patients who need urgent medical resources and special care. More importantly, this DL system is fast and does not require human-assisted image annotation, which increases its clinical value and become more robust. For a typical chest CT scan of a patient, the DL system takes less than ten seconds for prognostic and diagnostic prediction.

During building and training the DL system, we did not involve any human annotation to tell the system where the inflammatory area was. However, the DL system managed to automatically discover the important features that are strongly associated with COVID-19. In figure 5, we visualized the DL-discovered suspicious lung areas that were used by the DL system for inference. These DL-discovered suspicious lung areas have high overlap with the actual inflammatory areas that are used by radiologists for diagnosis. In previous studies, some radiological features such as GGO, crazy-paving pattern, and bilateral involvement are reported to be important for diagnosing CVOID-19 [7]. In the DL-discovered suspicious lung areas, we also observed these radiological features. This demonstrates that the high-dimensional features mined by the DL system can probably reflect these reported radiological finding.

Despite the good performance of the DL system, this study has several limitations. First, there are other prognostic end events such as death or admission to intensive care unit, and they were not considered in this study. Second, the management of severe and mild COVID-19 are different, thereby, explore prognosis of COVID-19 in these two groups separately should be helpful.

### Contributors

YZ and JT conceived and designed the study. SW implemented the DL system and wrote the paper. QW, YZ, and LW contributed to the data process and analysis. MN, HY, WG, YB, XQ, LL, XL, MW, HL, and WL contributed to data collection.

## Data Availability

Please contact the authors for data availability

## Acknowledgments

This paper is supported by the National Natural Science Foundation of China under Grant Nos. 81930053 and 81227901, the National Key R&D Program of China under Grant Nos. 2017YFA0205200. We thank all the colleagues in CAS key laboratory of Molecular Imaging, and collaborative hospitals for data collection.

